# Validation and implementation of a mobile app decision support system for quality assurance of tumor boards. Analyzing the concordance rates for prostate cancer from a multidisciplinary tumor board of a University Cancer Center

**DOI:** 10.1101/2022.05.27.22274488

**Authors:** Yasemin Ural, Thomas Elter, Yasemin Yilmaz, Michael Hallek, Rabi Raj Datta, Robert Kleinert, Axel Heidenreich, David A. Pfister

## Abstract

**Background:** Certified Cancer Centers must present all patients in multidisciplinary tumor boards (MTD), including standard cases with well-established treatment strategies. Too many standard cases can absorb much of the available time, which can be unfavorable for the discussion of complex cases. In any case, this leads to a high quantity, but not necessarily a high quality of tumor boards

**Objective:** Our aim was to develop a partially algorithm-driven decision support system (DSS) for smart phones to provide evidence-based recommendations for first-line therapy of common urological cancers. To assure quality, we compared each single digital decision with recommendations of an experienced MTD and obtained the concordance.

**Design, setting and participants:** 1873 prostate cancer patients presented in the MTD of the urological department of the University Hospital of Cologne from 2014 to 2018 have been evaluated.

**Outcome measurements and statistical analysis:** Patient characteristics included age, disease stage, Gleason Score, PSA and previous therapies. The questions addressed to MTD were again answered using DSS. All blinded pairs of answers were assessed for discrepancies by independent reviewers.

**Results and limitations:** Overall concordance rate was 99.1% (1856/1873). Stage specific concordance rates were 97.4% (stage I), 99.2% (stage II), 100% (stage III), and 99.2% (stage IV.Quality of concordance were independent of age and risk profile.

**Conclusions:** The reliability of any DSS is the key feature before implementation in clinical routine. Although our system appears to provide this safety, we are now performing cross-validation with several clinics to further increase decision quality and avoid potential clinic bias.

**Patient summary:** The quality of therapeutic decisions provided in tumor boards is perhaps the most relevant criterion for optimal cancer outcome. This tool aims to provide optimal recommendations, to assess the quality on a case-by-case basis and furthermore to objectively display the quality of oncological care.

**Author summary:** Everyday clinicians face the difficult task to choose the optimal treatment for their cancer patients due to the emergence of newly available therapeutics and continuously altering treatment guidelines. The resulting flood of information is impossible for clinicians to keep up with. Therefore, clinicians decide as a team, in so called tumor boards, upon the best possible cancer treatment for each patient. Even though the treatment decisions recommended by tumor boards play a critical role for the long-term survival of cancer patients, their accuracy in decision-making has hardly ever been assessed. Unfortunately, current digital tools that have been developed to support clinicians on the process of decision-making, have failed to provide treatment recommendations with sufficient accuracy. Therefore, we evaluated the quality of a novel decision-making application by comparing the decision concordance generated by the App with therapeutic recommendations given by a tumor board of a University Cancer Center. For newly diagnosed cancer patients we found that the novel tool matched the decisions made by the tumor board in almost 100% of the cases. These promising results not only show the potential providing digital support for patient care, but also provide objective quality management while saving board time in favor of discussing more complex cases.

## Introduction

Uro-Oncologists and Oncologists worldwide face the challenge of ensuring that their patients receive the best possible, individualized care for their cancer disease. Keeping up with the fast developments in medicine is very difficult for many physicians, as the rapid growth of scientific knowledge leads to an almost unmanageable variety of new treatment options [1]. The large amount of data is overwhelming and consequently it becomes a demanding task, to decide for the best possible individualized therapy for the patient. Therefore, case discussion in multidisciplinary tumor board (MTB) conferences is one of the most important factors to assure highest quality standards in oncological care. Driven by this assumption, German hospitals are required to present and discuss each cancer patient in MTB in order to get registered as certified cancer center by the German Cancer Society (Deutsche Krebshilfe) [2].

In clinical reality, certification requirements for presenting all routine cases leads to a significant increase in the number of case discussions, wasting valuable time and attention that is needed for the discussion of more complex tumor cases.

In the field of evidence-based oncology, it is almost paradoxical that the quality of individual therapeutic decisions of the tumor boards is hardly ever qualitatively assessed. Despite their widespread use in clinical routine, few data is available about the effects of tumor boards on quality of care and long-term survival of cancer patients [3,4].

This results in the need to objectively define and measure the quality of MTB at the level of individual therapeutic recommendations.

Without doubt, clinical decision support systems will play a major role in the near future, including artificial intelligence (AI) based systems, to close the gap between complex data and clinical decision-making [4-6]. Up to this point, systems based on artificial intelligence have been unable to offer reliable assistance in this area, as they are still failing to provide treatment recommendations with sufficient certainty even to standard questions on first-line therapy [7]. For example, one of the leading AI-based systems, Watson for Oncology, matched only 12% (stomach), 80% (colon and breast carcinoma) and 93% (ovarian carcinoma) of the treatment recommendations given by medical experts [8-13].

Another approach to provide digital therapeutic recommendations with sufficient certainty could be implemented by developing software based on clinical network expertise. This concept of expert-curated digital decision support systems (DSS) was described in Nature Biotechnology in 2018 and a comparison with approaches of artificial intelligence showed multiple benefits [14]. The main advantage described here is certainly that the expert systems seem to represent clinical reality better than the AI-based systems used so far.

The DSS smartphone application EasyOncology, whose digital treatment recommendations are based on continuous matching with real tumor boards, follows this approach and led to the design of this research study.

The aim of this clinical research is to implement the aforementioned technology for validation and quality assurance of a urological tumor board.

## Materials and methods

### Smartphone application

The content of EasyOncology was developed by experienced specialists from all relevant oncological specialties with the intention to provide evidence-based and unbiased first-line treatment recommendations for the most common cancer entities.

The intuitive user interface and quality of the application led to a top 3 ranking in a worldwide comparison of 157 oncological applications in 2017 [15].

The content and therapeutic recommendations are based on expert-curated knowledge that is assimilated and interpreted from written and visual sources alike. More precisely, given oncological guidelines (i.e. S3-guidelines, NCCN), approval status of therapeutics and best-clinical practice of major cancer centers build the basis for EO’s data platform.

Query algorithms have been developed to effectively support the decision-making process, especially in complex cancer cases. EasyOncology was certified as a medical device in July 2020. To assure quality of given treatment recommendations, a continuous comparison with real-word treatment recommendations of various MTDs has been implemented.

For this study, clinicopathological data of an uro-oncological MTD, namely the patient characteristics age, cancer stage, risk stratification, Gleason Score, and PSA-level (prostate specific antigen), were assessed to provide treatment recommendations using EasyOncology.

### Study Design and Patients

The study obtained ethics approval by the Ethics Commission of Cologne University’s Faculty of Medicine.

We present the results of 1873 cases of prostate carcinoma. We compared and analyzed the concordance rates between the tumor board recommendations of the urological multidisciplinary tumor board at the University of Cologne and the query results of the digital application “EasyOncology”.

Data sets of 2412 prostate cancer cases presented at university uro-oncology MTB during the time period of 2014-2018 were screened for inclusion criteria. 140 case discussions not addressing therapeutic procedures, such as specific questions about histopathology or how to obtain a biopsy were excluded, as well as another 264 cases without therapeutic decisions due to pending clinical information. Another 135 cases recommended for clinical trials (n=50) or complex cases with more than one active tumor entity (n=85) were also excluded (Fig 1).

**Fig 1.**
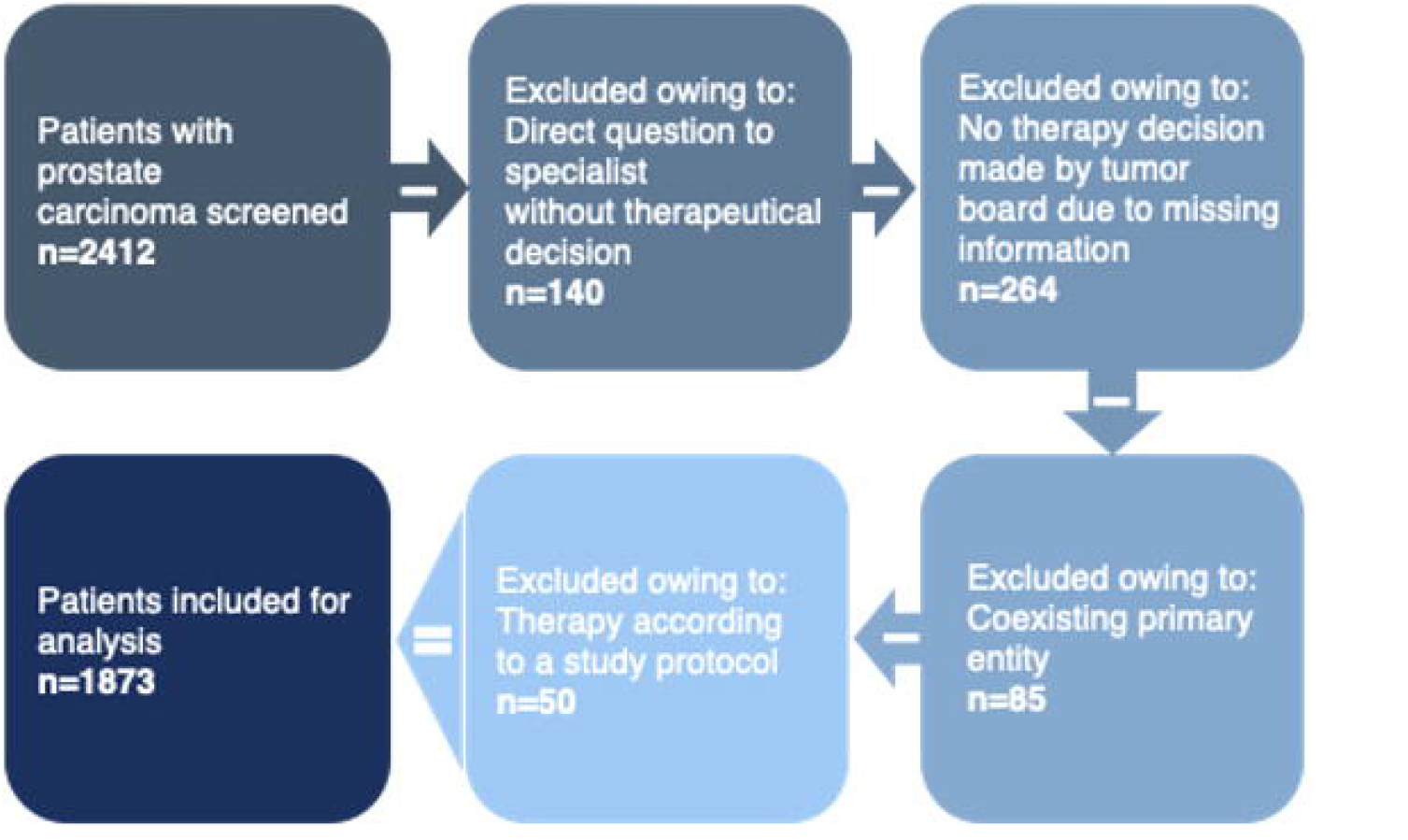
Flow chart patient cohort with prostate carcinoma.

The real-world and digital treatment recommendations given for each tumor board question were compiled as response pairs and blinded to their origin. Subsequently, the response pairs were first examined for agreement and responses that were not obviously identical.

The similarities and deviations of each pair of answers were evaluated by independent uro-oncology specialists and reported according to previous publications on Watson for Oncology by IBM (Fig 2) [9-12,16]:

**Fig 2.**
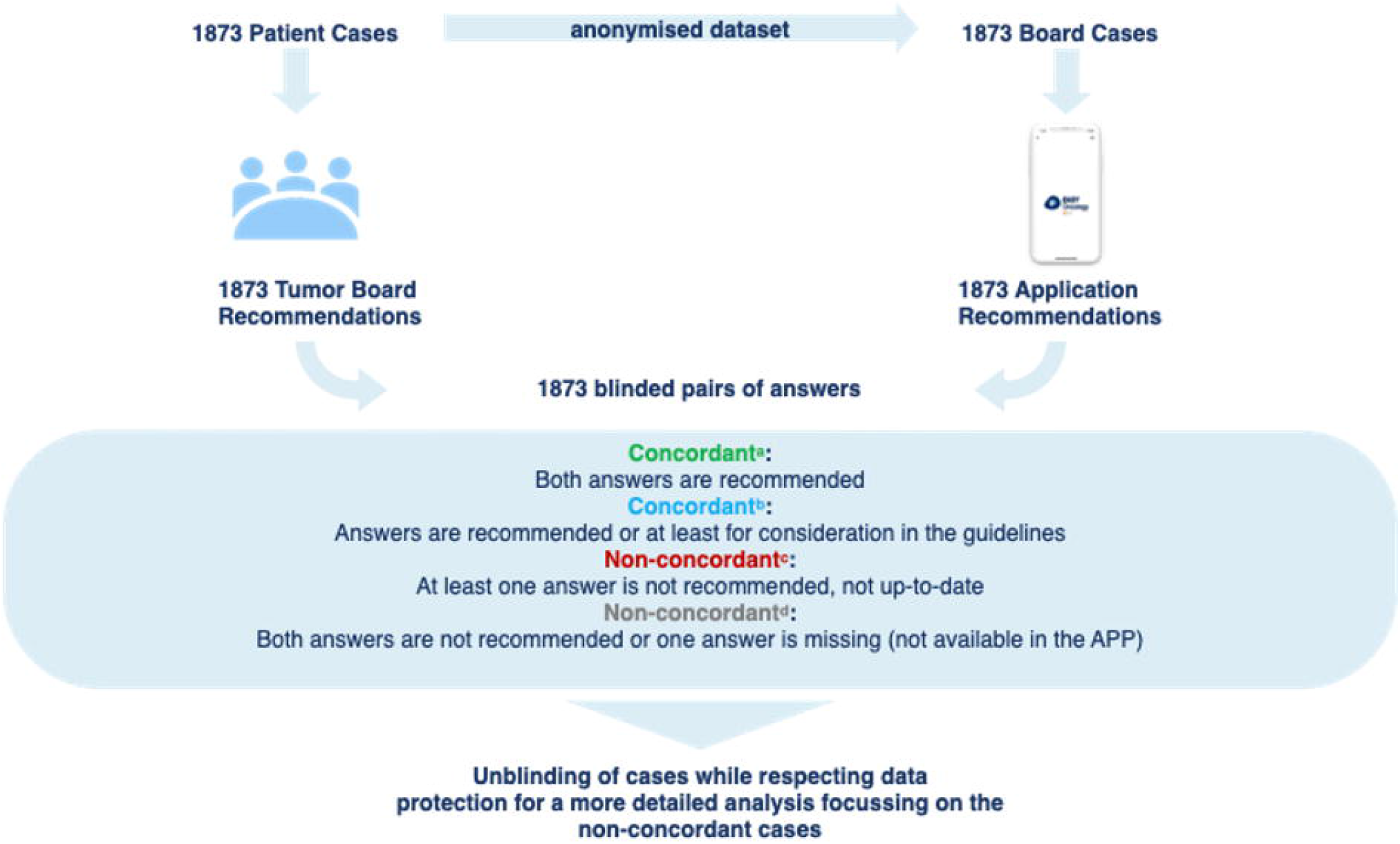
Evaluation flow chart.

1. “concordant, recommended” if both recommendations of the DSS application and MTD were identical
2. At first evaluation non-concordant cases, that were reviewed by independent specialist and judged as correct alternative treatment option were considered as “concordant, for consideration”
3. Case pairs were classified as “non-concordant, not recommended” if one of the recommendations, either App or MTD, did not meet current best-clinical-practice treatment guidelines
4. Pairs of cases were “non-concordant, not available” if the DSS could not provide a treatment recommendation due to missing information that is needed by the APP to give a treatment recommendation

Testing results were categorized into 4 color-coded groups: green^a^ represents “concordant treatment recommendation”; blue^b^ represents “concordant, for consideration”; red^c^ represents “non-concordant, not recommended” and grey^d^ represents “non-concordant, not available” recommendations.

In the second round of analysis, the mismatched pairs of responses were reviewed in detail in order to identify limitations in the query algorithm leading to non-concordancy and, subsequently, to improve the query.

### Data analysis and statistics

Descriptive statistics and data analysis were carried out using IBM’
ss statistics software SPSS Version 25 and Microsoft Excel. The patient characteristics age, cancer stage, risk stratification, Gleason Score, and PSA-level (prostate specific antigen) were documented.

Descriptive statistics were depicted as number of percentages or mean ± standard deviation (SD). After assigning patients to the concordant or the non-concordant group, a chi-squared test was used to compare categorical variables and the Mann-Whitney U test was applied to compare ordinal variables between the groups. Multivariate logistic regression analysis was used to analyze the association between the concordance rate and clinicopathological data. Statistical significance was assumed if the p-value was < 0.05 for all statistical analysis. Graphics, charts and tables were generated using SPSS, Microsoft Excel and Power Point.

## Results

The mean age of all patients was 68 years. Regarding stage classification, 238 (13%) of cases were classified as stage I, 519 (28%) as stage II, 262 (14%) as stage III, and 848 (45%) as stage IV. Of the 776 cases with localized disease, 331 cases (43%) presented with good prognosis, 330 (42%) with intermediate prognosis, and 115 cases (15%) with poor prognosis according to D’
sAmico classification.

Cases were categorized according to clinical status as non-metastatic hormone-naïve and treatment-naïve prostate cancer (46%); metastatic hormone-naïve prostate cancer (11%); and castrate resistant prostate cancer (19%). A further subset included cases dealing with follow-up; or treatment options after radical prostatectomy (RPE) with a histological R1 or pN1 situation; local or biochemical relapses; or questions regarding transurethral or suprapubic resection of the prostate (TUR-P/SPE) after incidental detection of prostate cancer (24%).

Using multivariate analysis, the Gleason score and the prognostic Grade (ISUP) were significantly associated with concordance rate (p=.001 each). This analysis and other patient characteristics are summarized in Table 1.

**Table 1.**
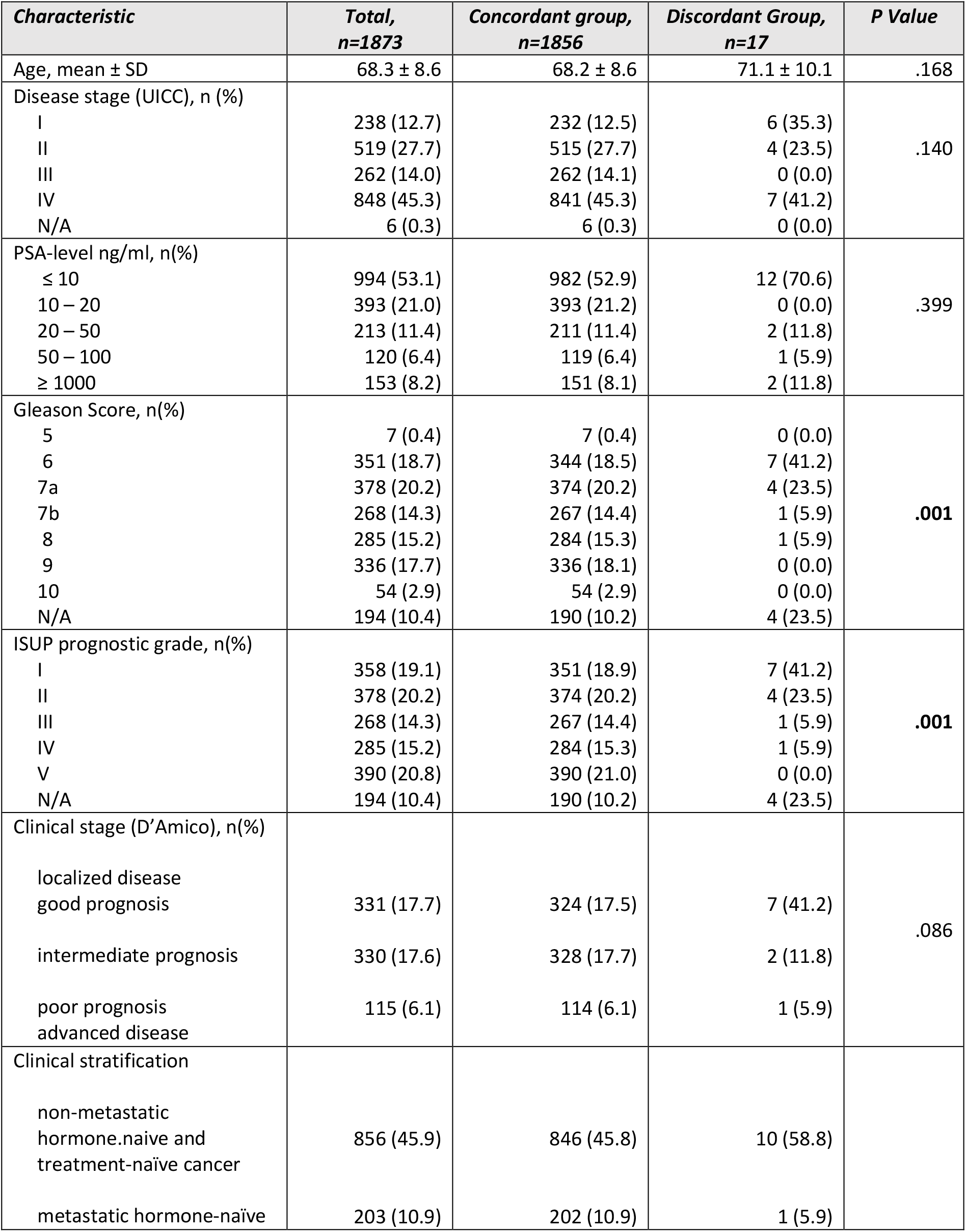
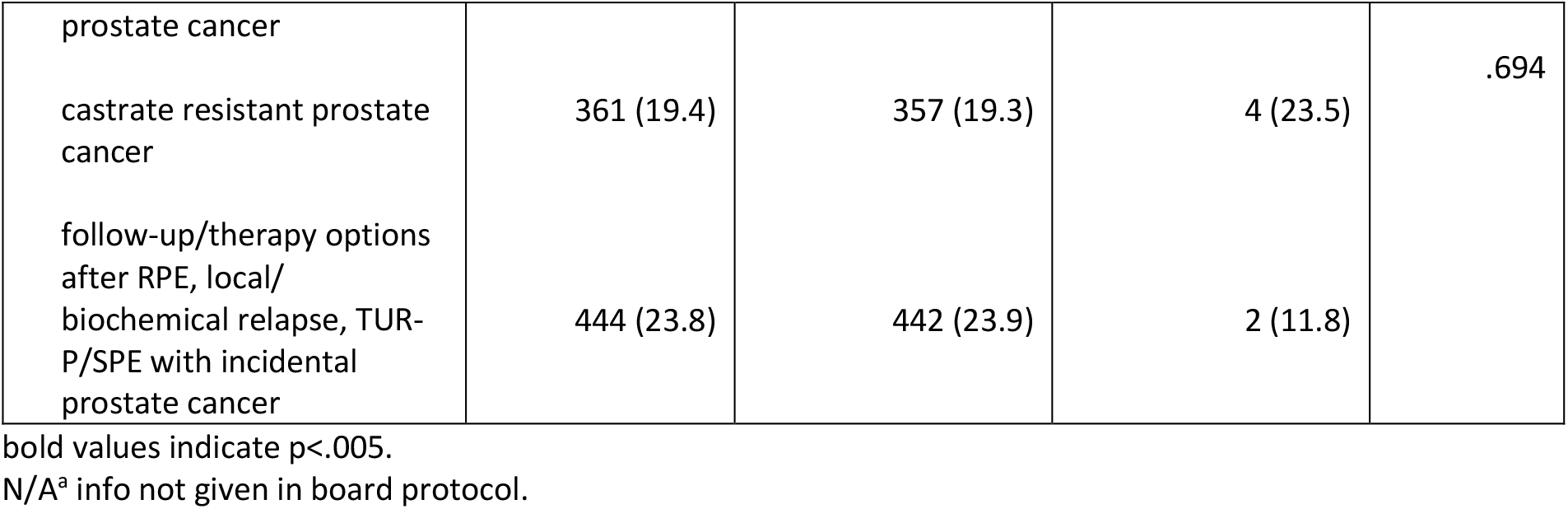
Baseline clinical characteristics.

The overall concordance rate between the actual treatments received by patients and cancer treatment recommendations given by EasyOncology for prostate cancer was 99%. Fig 3 shows the overall concordance rate.

**Fig 3.**
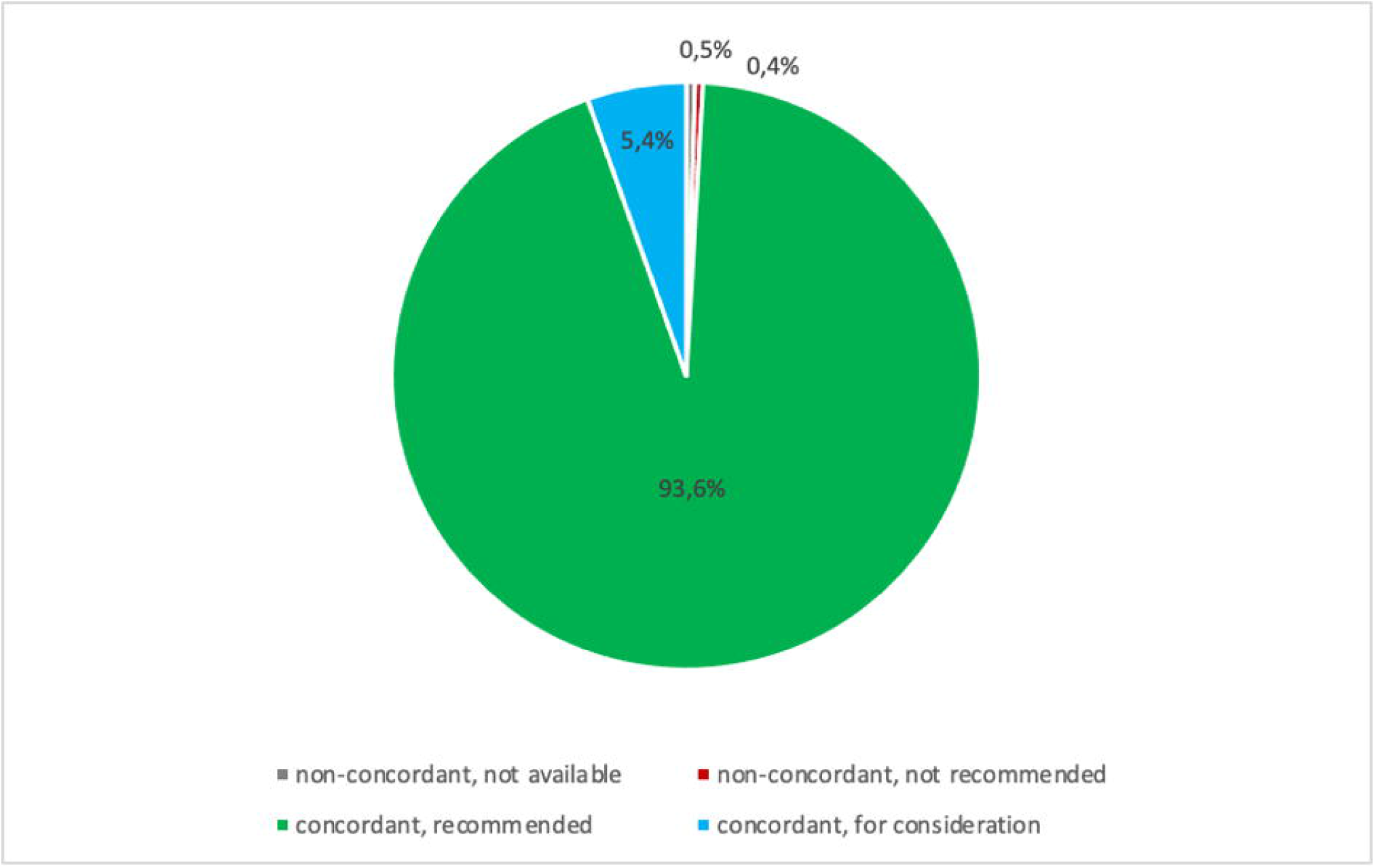
Overall treatment concordance between a multidisciplinary tumor board and the application EasyOncology.

As illustrated in Fig 4, stage specific concordance rates were 97.4% (stage I), 99.2% (stage II), 100% (stage III), and 99.2% (stage IV). Quality of concordance was independent of age, stage of disease and risk profile. The treatment concordance rates by age for > 50 years, 50 - 60 years, 60 - 70 years, 70 - 80 years, and ≥ 80 years were 100%, 99.0%, 99.6%, 99.0% and 97.0%, respectively. Patients with stage III cancer or who were <50 years old showed the highest concordance rates (100%).

**Fig 4.**
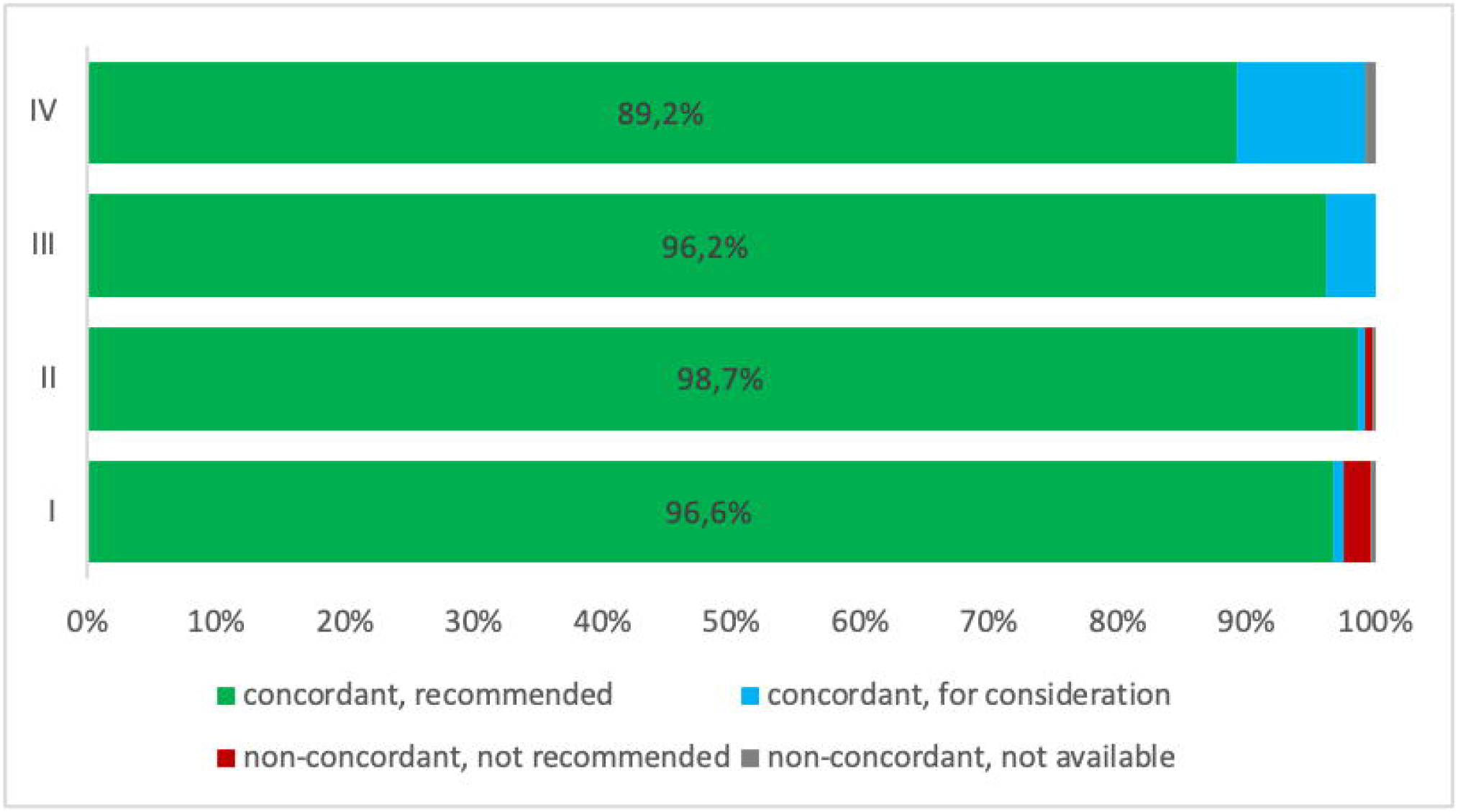
Concordance rates by Tumor Stage.

Overall, non-concordant results were found in 17 cases (Fig 4). As requested by protocol, all non-concordant cases were reviewed by an independent uro-oncological specialist for exact sub-classification of non-matching cases.

After review, eight of these cases were classified as “non-concordant, not recommended”; nine cases as “non-concordant, not available”.

Exemplary for a result that was rated “non-concordant, not recommended” to the disadvantage of the MTD was the case of a patient with newly diagnosed localized prostate cancer (UICC stage IIIA). Since the patient had a high PSA-level, the application recommended surgery or radiation, whereas MTD decided for active surveillance. The correct recommendation of the APP according to guidelines was confirmed by the reviewer. Nevertheless, MTD decision followed patient’s request for non-intervention.

Another similar example of a “non-concordant, not recommended” case for independent review was a patient with localized prostate cancer (UICC stage I). The application recommended a therapeutic intervention in accordance with current guidelines, as two positive biopsies were documented in the board protocol [17,18]. Here again, individual decision (patient request) led to the MTD recommendation for active surveillance.

In the remaining six “non-concordant, not recommended” cases, the DSS recommended an active therapy based on the information that more than two biopsies were positive, indicating a higher-risk disease. MTD attending specialists realized that these biopsies were obtained only from one single tumor lesion, which is not fulfilling high-risk criteria, and therefore correctly dismissed the idea of an active therapy in favor of active surveillance. Nine “non-concordant, unavailable” cases were identified as stage III neuroendocrine carcinomas, a histologic subtype that is not thematically considered by the APP and thus, no therapeutic recommendation was provided by DSS.

The queries of the APP on how many biopsy specimens were obtained, the patient’s wish against any active therapy and the presence of a neuroendocrine tumor could thus be identified as systematic errors for divergent treatment recommendations. These valuable insights can be used to optimize the APP in order to increase the reliability of its recommendations.

Using multivariate logistic regression analysis with independent variables age, PSA value and stage of disease (I/II vs. III/IV) no variables were found to be significantly associated with a decrease in the concordance rate. The results of the multivariate logistic regression are detailed in Table 2.

**Table 2.** Multivariate regression analyses of the concordance rate between EasyOncology and the multidisciplinary tumor board.

| **Variables** | **Multivariate Analysis** |  |  |
| --- | --- | --- | --- |
|  | **OR** | **95% CI** | **P value** |
| Age | .955 | .898 – 1.015 | .141 |
| PSA (ng/ml) | 1.000 | 1.000 – 1.001 | .985 |
| Stage (≥3) | 2.210 | .832 – 5.870 | .112 |

## Discussion

The rapid development of new, innovative oncological treatment options leads more than ever to the requirement of quality-assured therapeutic decisions [17]. In order to give optimal treatment recommendations, physicians usually follow guidelines of medical societies, inform themselves through professional journals, participate in congresses, further their medical education and discuss cases in multidisciplinary tumor boards (MTB).

However, who can guarantee that all doctors working in oncology have the time and motivation to handle the information overload? How do they deal with this situation when even current guidelines of the medical associations sometimes fail to mention highly effective and newly approved therapeutics? Who ensures that the expertise of the doctors attending the MTB is actually given and that decisions are not (consciously or unconsciously) influenced by economic motives? Is there any evidence at all that tumor boards generally improve the quality of oncology care? [3,17-19]

AI-based systems seem to have the potential to support clinical decision making, as they have already impressively demonstrated their outstanding superiority in medical image processing and interpretation for different cancer entities [20-23]. Especially since AI-based systems seem perfectly suited to capture and correlate the immense amount of oncological knowledge, the results of all relevant clinical trials and all published case reports, and, based on this knowledge, to finally generate therapeutic recommendations. As obvious as this sounds, it is almost surprising that AI-based systems have yet failed to establish themselves in clinical oncological routine. So far, all attempts of AI-systems to reliably provide even standard therapeutic recommendations for first-line therapy have been disappointing. For example, Watson for Oncology, the leading application in this field, showed a concordance rate of only 73.6% compared to recommendations made by medical professionals for the first-line therapy of prostate cancer [16]. Watson for Oncology also obtained comparatively low concordance rates in other tumor entities, such as 12% in gastric cancer [13,24], 46.4% to 65.8% in colon cancer [10,11] or 77% in differentiated thyroid carcinoma [25] and others [9,26,27]. When considering the implementation of AI systems, the framework provided by the existing healthcare system must be carefully considered.

The effort required to implement systems such as Watson for Oncology in hospitals is enormous. The use of AI systems requires data protection-compliant interoperability between many hospital information systems and the required data must be completely accessible in predefined files.

In Germany in particular, data protection requirements of 16 federal states and the non-standardized norms for data processing and storage pose considerable challenges for developers of AI systems. In addition, clinical data continues to be frequently stored in paper files and it should not be forgotten that the exchange of diagnostic reports between clinics, pathologists, oncologists and practices is to this day commonly carried out by a fax machine. Another point of criticism of AI systems is often that the decision-making process is not easy to understand and that one has to trust almost blindly in the correctness of the machine response. Furthermore, the validation efforts that are needed when using AI systems ties up considerable human resources since only medical experts can judge if the recommendations are correct.

Until these structural problems are solved, expert-curated solutions offer an alternative, as described in Nature Biotechnology in 2018 [14]. This approach was adopted by medical professionals using the DSS and is the basis of this research. In order to ensure the quality of the recommendations given by the application, a continuous comparison with tumor boards of certified cancer centers was implemented.

This comparably simple and resource-saving technical solution proved to be beneficial here, enabling the large number of tumor board recommendations to be effectively compared retrospectively.

For prostate carcinoma, our expert curated digital decision support system provides an optimal concordance rate with the therapeutic recommendations of a university tumor board.

Yet, the very high concordance rate of our system is probably not surprising, since we evaluated predominantly first-line cases, for which guidelines generally apply. Of course, the degree of complexity increases with each tumor recurrence and additional concomitant diseases.

However, this is exactly the specification of our work, which aims to reduce the workload of tumor boards by providing digitalized answers to non-complex routine cases.

Despite the fact that this approach achieves better results than other methods published so far, further limitations have to be taken into account.

It should be stated as a limiting factor of our work that a high concordance rate is easier to achieve when therapeutic strategies do not show a significant change over a longer period of time, as given during the time period studied, from 2014 to 2018 [28].

The increasing dynamics of diagnostic and therapeutic options in the treatment of prostate cancer thus leads to significantly more frequent and shorter testing intervals of the application, which has been certified as a medical device in the meantime, and to continuous adaptation to best-clinical-practice.

By continuously comparing digital and analog recommendations, systemic deficits that lead to deviations usually become quickly apparent, thus enabling the prompt adaptation of the query logic to the dynamic development of therapeutic options.

Second limiting bias, a 100% concordance rate is of little value if the recommendation quality of the reference board is not validated. This leads to the necessity of establishing decision networks in order to generate a recommendation basis that is as reliable as possible and provides the basis for the required safety of recommendations. Therefore, cross-validation of different urological cancer centers is ongoing in order to eliminate the single-center bias.

The main goal of this development is to provide reliable recommendations for standard cases in advance of the tumor board conference with the aim of allowing more time for the discussion of complex cases. It is not in the developers’ interest to achieve 100% agreement between tumor board responses and digital recommendations, as no automated system will be able to consider all the complex clinical circumstances.

Rather, a trusted DSS must be able to reliably identify complex clinical constellations, which should then be discussed by experts attending the tumor board. Particularly in the case of complex diseases, medical expertise is irreplaceable and must remain so. However, expertise requires time, and this time should not be spent discussing universally accepted standard procedures.

## Perspective options

It is almost paradoxical in evidence-based driven oncology that the actually relevant quality of individual therapeutic decisions is virtually unknown.

The use of intelligent software could ensure the quality of treatment on a case-by-case basis and thus serve as an instrument for quality assurance that can be transparently accessed and compare the quality of oncological care provided by hospitals and medical practices. Based on the smartphone application used in this work for recommendation matching, we developed an interface that enables the necessary inputs in the decision process of tumor boards. Easily integrated into any system, this validated and reliable application could unburden tumor boards from standard cases, thereby allowing more time for discussion of complex cases.

## Data Availability

All data contained in the manuscript as well as the primary metadata are based on the tumor board documentation of the University Hospital Cologne. The data were evaluated in a pseudonymized form in compliance with data protection guidelines and can be assigned to the real cases by the treating physicians who are permitted to inquire about them.

## References

1. Global Oncology Trends 2019: Therapeutics, Clinical Development and Health System Implications. (QVIA Institute, 2019).

2. Deutsche Krebsgesellschaft und Deutsche Krebshilfe. Nationales Zertifizierungsprogramm Krebs. Erhebungsbogen für Onkologische Spitzenzentren und Onkologische Zentren, Inkraftsetzung am 03.12.2019.

3. Keating, N.L. et al. Tumor boards and the quality of cancer care. J Natl Cancer Inst 105, 113–21 (2013).

4. Soukup, T., Lamb, B.W., Weigl, M., Green, J.S.A. & Sevdalis, N. An Integrated Literature Review of Time-on-Task Effects With a Pragmatic Framework for Understanding and Improving Decision-Making in Multidisciplinary Oncology Team Meetings. Front Psychol 10, 1245 (2019).

5. Chen, Y., Elenee Argentinis, J.D. & Weber, G. IBM Watson: How Cognitive Computing Can Be Applied to Big Data Challenges in Life Sciences Research. Clin Ther 38, 688–701 (2016).

6. Letzen, B., Wang, C.J. & Chapiro, J. The Role of Artificial Intelligence in Interventional Oncology: A Primer. J Vasc Interv Radiol 30, 38–41 e1 (2019).

7. Schmidt, C. M. D. Anderson Breaks With IBM Watson, Raising Questions About Artificial Intelligence in Oncology. J Natl Cancer Inst 109(2017).

8. Seidman, A.D. et al. Integration of multi-modality treatment planning for early stage breast cancer (BC) into Watson for Oncology, a Decision Support System: Seeing the forest and the trees. Journal of Clinical Oncology 33, e12042–e12042 (2015).

9. Somashekhar, S.P. et al. Watson for Oncology and breast cancer treatment recommendations: agreement with an expert multidisciplinary tumor board. Ann Oncol 29, 418–423 (2018).

10. Kim, E.J. et al. Early experience with Watson for oncology in Korean patients with colorectal cancer. PLoS One 14, e0213640 (2019).

11. Lee, W.S. et al. Assessing Concordance With Watson for Oncology, a Cognitive Computing Decision Support System for Colon Cancer Treatment in Korea. JCO Clin Cancer Inform 2, 1–8 (2018).

12. Zhou, N. et al. Concordance Study Between IBM Watson for Oncology and Clinical Practice for Patients with Cancer in China. Oncologist 24, 812–819 (2019).

13. Choi, Y.I. et al. Concordance Rate between Clinicians and Watson for Oncology among Patients with Advanced Gastric Cancer: Early, Real-World Experience in Korea. Can J Gastroenterol Hepatol 2019, 8072928 (2019).

14. Bungartz, K.D., Lalowski, K. & Elkin, S.K. Making the right calls in precision oncology. Nat Biotechnol 36, 692–696 (2018).

15. Calero, J.J., Oton, L.F. & Oton, C.A. Apps for Radiation Oncology. A Comprehensive Review. Transl Oncol 10, 108–114 (2017).

16. Yu, S.H. et al. Early experience with Watson for Oncology: a clinical decision-support system for prostate cancer treatment recommendations. World J Urol (2020).

17. Specchia, M.L. et al. The impact of tumor board on cancer care: evidence from an umbrella review. BMC Health Serv Res 20, 73 (2020).

18. Devitt, B., Philip, J. & McLachlan, S.A. Re: Tumor boards and the quality of cancer care. J Natl Cancer Inst 105, 1838 (2013).

19. Krasna, M., Freeman, R.K. & Petrelli, N.J. Re: Tumor boards and the quality of cancer care. J Natl Cancer Inst 105, 1839–40 (2013).

20. Esteva, A. et al. Dermatologist-level classification of skin cancer with deep neural networks. Nature 542, 115–118 (2017).

21. Ardila, D. et al. End-to-end lung cancer screening with three-dimensional deep learning on low-dose chest computed tomography. Nat Med 25, 954–961 (2019).

22. Rubin, R. Artificial Intelligence for Cervical Precancer Screening. JAMA 321, 734 (2019).

23. McKinney, S.M. et al. International evaluation of an AI system for breast cancer screening. Nature 577, 89–94 (2020).

24. Tian, Y. et al. Concordance Between Watson for Oncology and a Multidisciplinary Clinical Decision-Making Team for Gastric Cancer and the Prognostic Implications: Retrospective Study. J Med Internet Res 22, e14122 (2020).

25. Kim, M. et al. Concordance in postsurgical radioactive iodine therapy recommendations between Watson for Oncology and clinical practice in patients with differentiated thyroid carcinoma. Cancer 125, 2803–2809 (2019).

26. Liu, C. et al. Using Artificial Intelligence (Watson for Oncology) for Treatment Recommendations Amongst Chinese Patients with Lung Cancer: Feasibility Study. J Med Internet Res 20, e11087 (2018).

27. Zhou, N. et al. Concordance Study Between IBM Watson for Oncology and Clinical Practice for Patients with Cancer in China. Oncologist (2018).

28. Interdisziplinäre Leitlinie der Qualität S3 zur Früherkennung, Diagnose und Therapie der verschiedenen Stadien des Prostatakarzinoms Version 5.0 April 2018

